# Whole exome-seq and RNA-seq data reveal unique neoantigen profiles in Kenyan breast cancer patients

**DOI:** 10.1101/2024.06.18.24309133

**Authors:** Godfrey Wagutu, John Gitau, Kennedy Mwangi, Mary Murithi, Elias Melly, Alexandra R. Harris, Shahin Sayed, Stefan Ambs, Francis Makokha

## Abstract

**Background:** The immune response against tumors relies on distinguishing between self and non-self, the basis of cancer immunotherapy. Neoantigens from somatic mutations are central to many immunotherapeutic strategies and understanding their landscape in breast cancer is crucial for targeted interventions. We aimed to profile neoantigens in Kenyan breast cancer patients using genomic DNA and total RNA from paired tumor and adjacent non-cancerous tissue samples of 23 patients.

**Methods:** We sequenced the genome-wide exome (WES) and RNA, from which somatic mutations were identified and their expression quantified, respectively. Neoantigen prediction focused on human leukocyte antigens (HLA) crucial to cancer, HLA type I. HLA alleles were predicted from WES data covering the adjacent non-cancerous tissue samples, identifying four alleles that were present in at least 50% of the patients. Neoantigens were deemed potentially immunogenic if their predicted median IC50 binding scores were ≤500nM and were expressed [transcripts per million (TPM) >1] in tumor samples.

**Results:** An average of 1465 neoantigens covering 10260 genes had ≤500nM median IC50 binding score and >1 TPM in the 23 patients and their presence significantly correlated with the somatic mutations (*R*^2^ =0.570, *P*=0.001). Assessing 58 genes reported in the catalog of somatic mutations in cancer (COSMIC, v99) to be commonly mutated in breast cancer, 44 (76%) produced >2 neoantigens among the 23 patients, with a mean of 10.5 ranging from 2 to 93. For the 44 genes, a total of 477 putative neoantigens were identified, predominantly derived from missense mutations (88%), indels (6%), and frameshift mutations (6%). Notably, 78% of the putative breast cancer neoantigens were patient-specific. HLA-C*06:01 allele was associated with the majority of neoantigens (194), followed by HLA-A*30:01 (131), HLA-A*02:01 (103), and HLA-B*58:01 (49). Among the genes of interest that produced putative neoantigens were *MUC17*, *TTN*, *MUC16*, *AKAP9*, *NEB*, *RP1L1*, *CDH23*, *PCDHB10*, *BRCA2*, *TP53*, *TG*, and *RB1*.

**Conclusions:** The unique neoantigen profiles in our patient group highlight the potential of immunotherapy in personalized breast cancer treatment as well as potential biomarkers for prognosis. The unique mutations producing these neoantigens, compared to other populations, provide an opportunity for validation in a much larger sample cohort.

## INTRODUCTION

Breast cancer is among the most frequent causes of cancer-related mortality in women. Disease heterogeneity and limited immunogenicity contribute to the lethality of breast cancer (Benvenuto et al., 2019). Immune evasion, an important hallmark of cancer, adds to the complexity of cancer burden through induction of immunosuppression (Bates et al., 2018). Immune checkpoint blockade (CKB) therapy has been developed to target and block immune regulatory molecules (PD-1/PD-L1 and CTLA-4) and in the process reactivate T cell immunity (Touchaei &Vahidi, 2024). This approach has been reported to improve clinical responses and survival, especially in tumors with high mutational burdens, such as lung cancer and melanoma (Shiravand et al., 2022). However, CKB therapy is not universally successful among all patients and shows increased efficacy with higher mutational burden tumors (Brahmer et al., 2015). Another immunotherapy approach that has been tested in clinical studies is the targeting of tumor-associated antigens (TAAs) that are expressed in tumors at abnormally high levels and rarely detectable in normal tissues (Valilou & Rezaei, 2019). One of the limitations of this therapy approach is that many TAAs represent normal self-antigens and thus can be tolerated by T-cells, resulting in poor immune response (Benvenuto et al., 2019). This poses challenges for applicability in breast cancer because it generally has a lower mutational burden. Thus, CKB and TAAs immunotherapy have had limited success in breast cancer patients (Narang et al., 2019).

Tumor neoantigens are tumor-specific antigens derived from somatic mutations in expressed genes and are presentable to the major histocompatibility complex (MHC) by both class I human leukocyte antigen (HLA-I) molecules present on surface of cancer cell, as well as class II HLA molecules present on professional antigen-presenting cells (Blass & Ott, 2021). This elicits anti-tumor immune responses that have the potential of eliminating the tumor cells with minimal off-target effects (Pan et al., 2018). Neoantigens are encoded in various mutational types, including single nucleotide substitution, insertion and deletions (INDELs), splice sites, stop codons gains and silent change, which can result in translational frameshifts or novel open reading frames (Benvenuto et al., 2019). As such, these neoantigens offer an advantage over TAAs in that they are only expressed by cancer cells and not by normal cells, which enables specific recognition by the immune system (Benvenuto et al., 2019). Although some neoantigens are shared among patients, most of them are patient-specific and are not subject to immune tolerance mechanisms (Yarchoan et al., 2017). The specificity of neoantigens could provide an opportunity for future personalized therapy in a cancer with a low tumor mutational burden and a high disease heterogeneity, such as breast cancer. Moreover, neoantigens can potentially be used as biomarkers in cancer immunotherapy to assess or predict the response of a patient to treatment (Benvenuto et al., 2019).

Despite advancements in next generation sequencing and high-performance computing that has resulted in improved cancer immunotherapy research and neoantigen-based treatments, there remains a scarcity of information regarding neoantigens in specific populations from sub-Saharan African countries such as Kenya. This lack of data poses a significant challenge in tailoring immunotherapeutic strategies for breast cancer patients in such regions that have a high cancer burden, especially when compounded by germline ancestral factors and a distinct mutational spectrum that may influence tumor biology and immune response. Thus, it is critical to profile the neoantigen burden in this population to contribute to the global collection of breast cancer immunogenic antigens for future drug development. To this end, we sought to profile neoantigens in Kenyan women diagnosed with breast cancer *in silico* through analysis of the whole exome and RNA sequencing data from 23 patients. We characterized the mutation burden for each patient using WES, identified gene expression patterns in tumor tissue, and predicted the putative neoantigens incorporating these datasets.

## MATERIALS AND METHODS

### Patients and samples

Tumor and adjacent normal tissue pairs were obtained from 23 breast cancer patients at the Aga Khan Hospital, Nairobi, Kenya and AIC Kijabe Hospital, Kijabe, Kenya between 2019 and 2021. Samples were collected through surgical excision, after which tissues were snap frozen in liquid nitrogen and temporarily stored at Aga Khan Hospital. Frozen tissue samples were shipped to the National Cancer Institute, Bethesda, MD, USA, for sequencing. Prior to tissue collection, all patients provided written informed consent and the study was approved by Research and Ethics Committees at Aga Khan University Hospital, Nairobi (Ref: 2018/REC-80) and AIC Kijabe Hospital (KH IERC-02718/2019).

### Whole-exome sequencing (WES) and RNA-sequencing

Genomic DNA was extracted from the samples using the DNeasy Blood and Tissue Kit (Qiagen, Hilden, Germany), following manufacturer’s instructions. Total RNA was extracted from the frozen tissues using TRIzol reagent (Invitrogen). WES was performed by the company, Psomagen (https://www.psomagen.com/). This service provider is Clinical Laboratory Improvement Amendments-certified and College of American Pathologists (CAP)-accredited, achieving a sequence depth of 250x for tumor tissues and 150x for adjacent non-cancerous tissues, as previously described by us (Tang et al., 2023). Total RNA from the 23 sample pairs was processed by a NCI Leidos core facility, where library preparation was performed using the TruSeq Poly A kit (Illumina, San Diego, USA). Samples were sequenced on a Novaseq system with 150 bp paired-end reads and a depth of 30 million reads.

### Reads mapping and variant calling

For WES, raw reads were quality checked using FASTQC (Andrews, 2010) and results summarized using MultiQC (Ewels et al., 2016). The reads were trimmed for low quality reads and adapter sequences using Trimmomatic (Bolger et al., 2014) and quality-checked again using FASTQC and MultiQC. All samples passed the QC test after trimming and the reads were aligned using BWA-MEM (Li, 2013) to the hg38 human reference genome, where >95% of the reads aligned properly to the genome. The aligned reads were deduplicated and read groups added to the deduplicated bam files using Picard. This was followed by base quality recalibration in GATK (McKenna et al., 2010). Somatic variant calling was performed using MuTect2 (McKenna et al., 2010) in paired tumor-normal mode utilizing the panel of normal option that was derived from normal reads. Variants were normalized using a variant tool set (vt; Tan et al., 2015), filtered using GATK and functional/consequence-annotated using a variant effect predictor (VEP; McLaren et al., 2016). Annotated variants were converted to MAF files using *vcf2maf* (Kandoth et al., 2020) and concatenated into a single file. The MAF files were imported into R package *maftools* (Mayakonda et al., 2018) for further processing.

For RNA-seq, a quality check was performed using FASTQC and MultiQC after which the reads were trimmed and quality checked again. All samples passed the quality check and the reads were pseudo-aligned to the hg38 reference genome using Kallisto aligner (Bray et al., 2016) with default settings to obtain count matrix. Alignment statistics showed that over >50% reads mapped uniquely to the genome. The raw counts were normalized into estimated Transcripts Per Million (TPM), and scaled using the average transcript length over samples and the library size by *tximport* (Soneson et al., 2016).

### Variant expression annotation

VCF files containing the variants were annotated for expression using the vcf-expression-annotator (https://github.com/griffithlab/VAtools) with default setting except for choosing the use of gene names instead of transcripts and thereby ignoring the Ensembl id version. The tool takes the output of Kallisto and adds the data contained in the file to the VEP annotated VCF’s INFO column. Each of the variant annotated gets its expression value (TPM) added to the annotation information and this is used to determine the level of variant expression during neoantigen filtering.

### Neoantigen prediction

Human leukocyte antigen (HLA) class I alleles (HLA a, b and c) were predicted from each patient’s normal sample exome-seq data using HLA-HD v.1.2.1 (Kawaguchi et al., 2017). Here, the putative HLA reads are aligned to an imputed library of full-length HLA alleles. Neoantigens were then predicted using pVACseq (Hundal et al, 2016) with MHCflurry, MHCnuggetsI, SMM, and SMMPMBEC algorithms and keeping the default parameters, except for turning off the VAF and coverage filters. Here, the neoepitopes that could bind to the patient-specific HLA alleles were predicted from the Immune Epitope Database (IEDB; Vita et al., 2019). This involved matching patient HLA type to the existing IEDB list keeping all amino acids with lengths for 9, 10 and 11-mers. Predicted epitopes were filtered to retain only those with high affinity (IC50 ≤ 500nM) and were expressed (transcripts per million, TPM>1) in tumor samples. The bioinformatic analysis workflow is outlined in Figure 1.

**Figure 1:**
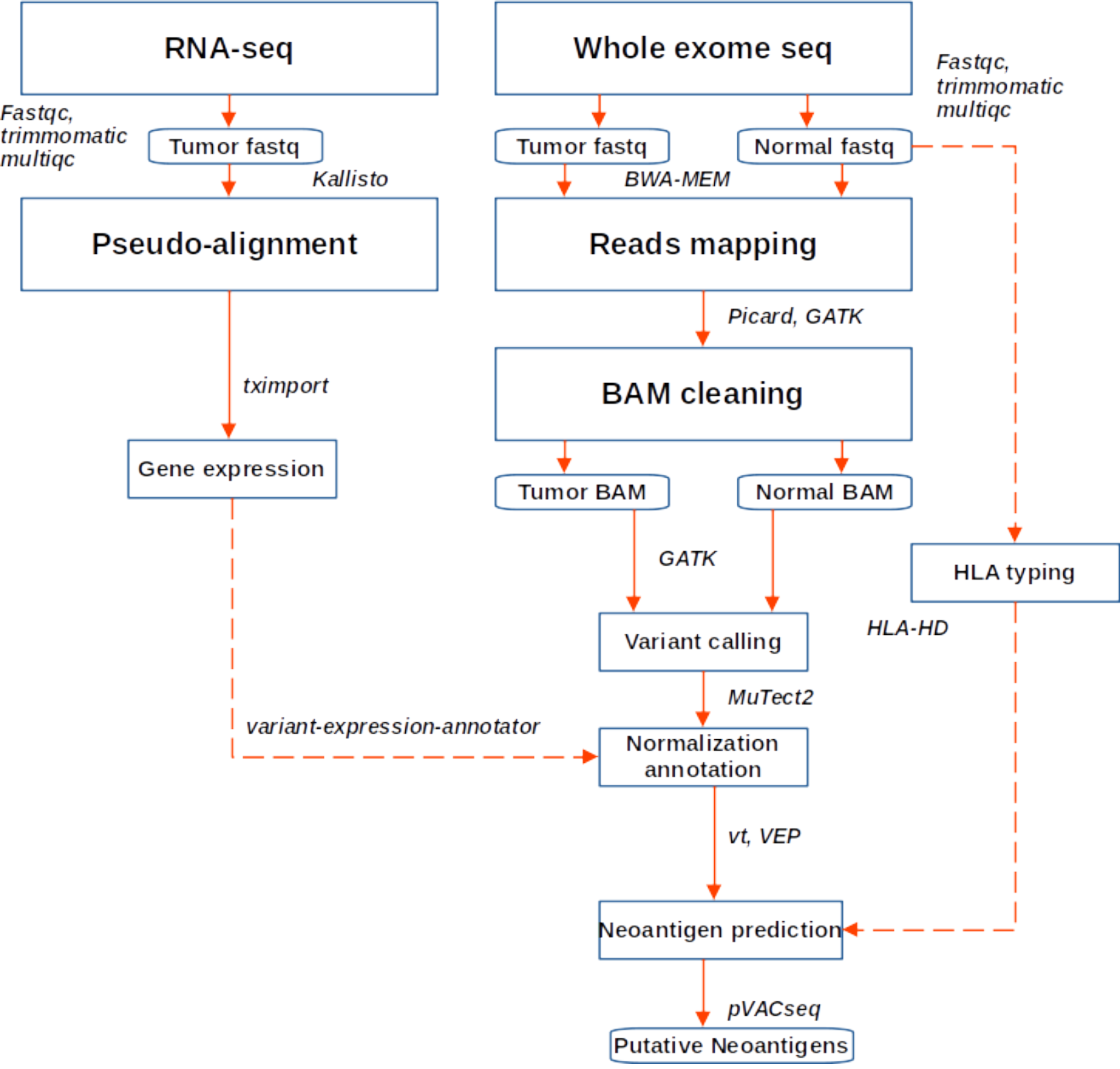
Workflow for neoantigen prediction from WES and RNA sequencing data. Fastq files were quality checked, trimmed and aligned to the hg38 genome. Variant calling was performed following GATK best practice, while gene expression was quantified using Kallisto. Variants were annotated and expression data added, after which neoantigen prediction was performed in PVACseq pipeline

Sample summary statistics and the pairwise tests for differences among mutations and neoantigens abundance among the BC subtypes using Wilcoxon test and visualization of the results were performed in R software (R Core Team, 2023).

## RESULTS

### Patients and sample characteristics

The demographic and clinical characteristics of the 23 breast cancer patients are summarized in supplementary Table S1. We grouped the tumors into 3 subtypes based on expression of either the hormone receptors (HR) or human epidermal growth factor receptor 2 (HER2) (Narang et al., 2019): those that were HER2+ regardless of the HR status, those that were negative for all hormone receptors (triple negative breast cancer; TNBC) and those that were HR+ and HER2-. Majority of the samples were HR+/HER2-constituting 52.2%, followed by HER2+ at 34.8% and TNBC at 13.0%. Most of the patients had invasive carcinoma (invasive ductal carcinoma, 78.26% and invasive carcinoma; 4.35%). For tumor grade, 65.22% of the patients had grade 3 tumors (65.22%), while the rest had grade 2 tumors (34.78%). Clinically, 39.13% of the patients were in stage II, 30.44% in stage III, and 8.7% in stage I (Table S1).

### Mutation profiles for the 23 patients

Across all genes, the average number of detected mutations in the 23 patients was 2809 mutations. Considering the different subtypes, TNBC had the highest average number of mutations at 3202, followed by HR+/HER2-at 2757, and HER2+ at 2740 mutations (Figure S1). From the catalog of somatic mutations in cancer (COSMIC, v99), we identified 73 genes reported to be mutated in breast cancer and among those, 62 (84.9%) had at least one mutation in our samples. The mutation frequency among the 62 genes ranged from 1 to 55 mutations per individual. The majority of the mutations were of the missense type, most of which were substitutions of C>T (Figure 2). The top 10 mutated genes among the 62 are shown in Figure 3. Four genes (*MUC16*, *MUC17*, *TTN*, *RP1L1*) were altered in more than 95% of the patients (Figure 3). Moreover, mutations in genes *TP53*-*ERBB3*, *PTEN*-*CFAP46* were found to significantly co-occur, while *BRCA1*-*MUC17* mutations were significantly mutually exclusive (*P*<0.05) (Figure 4). Furthermore, the majority of the single nucleotide mutations were substitution of C to T, whereas T to A substitutions were most uncommon. Transitions occurred more frequently than transversion in these substitutions (Figure 5).

**Figure 2:**
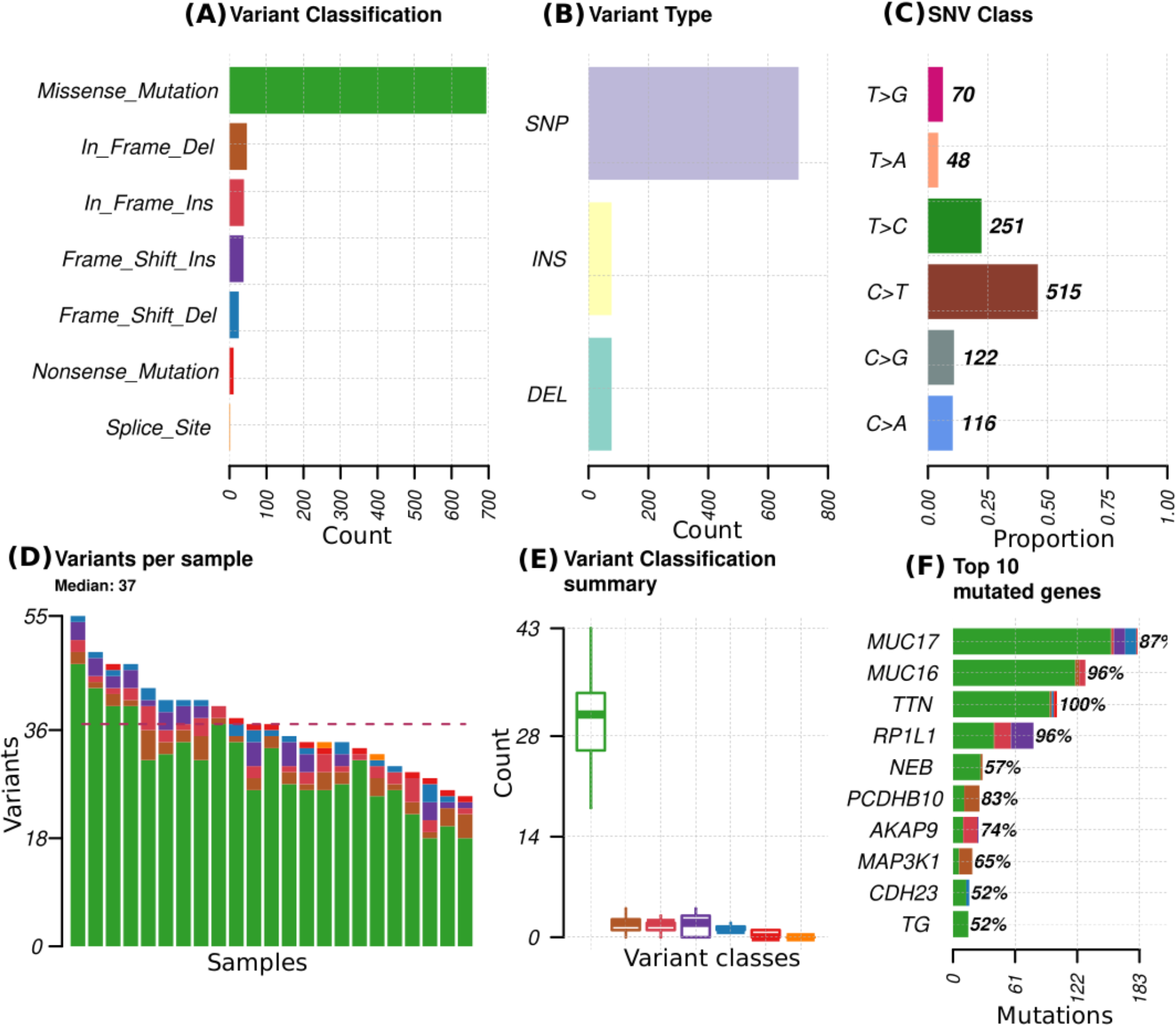
Mutational profiles in 23 patients for 73 genes reported to be mutated in breast cancer. **A**) variant classes abundance in the total mutations, **B**) variant types that include single nucleotide polymorphism (SNP), insertions (INS) and deletions (DEL), **C**) proportion of different single nucleotide variant (SNV), **D**) distribution of variants per sample with colors representing the different variant classes denoted in A, **E**) summary of the variant classes distribution and numbers in all samples, **F**) Top 10 mutated genes, with colors representing different variant classes and the percentages indicating the proportion of samples in which the genes mutations are present.

**Figure 3:**
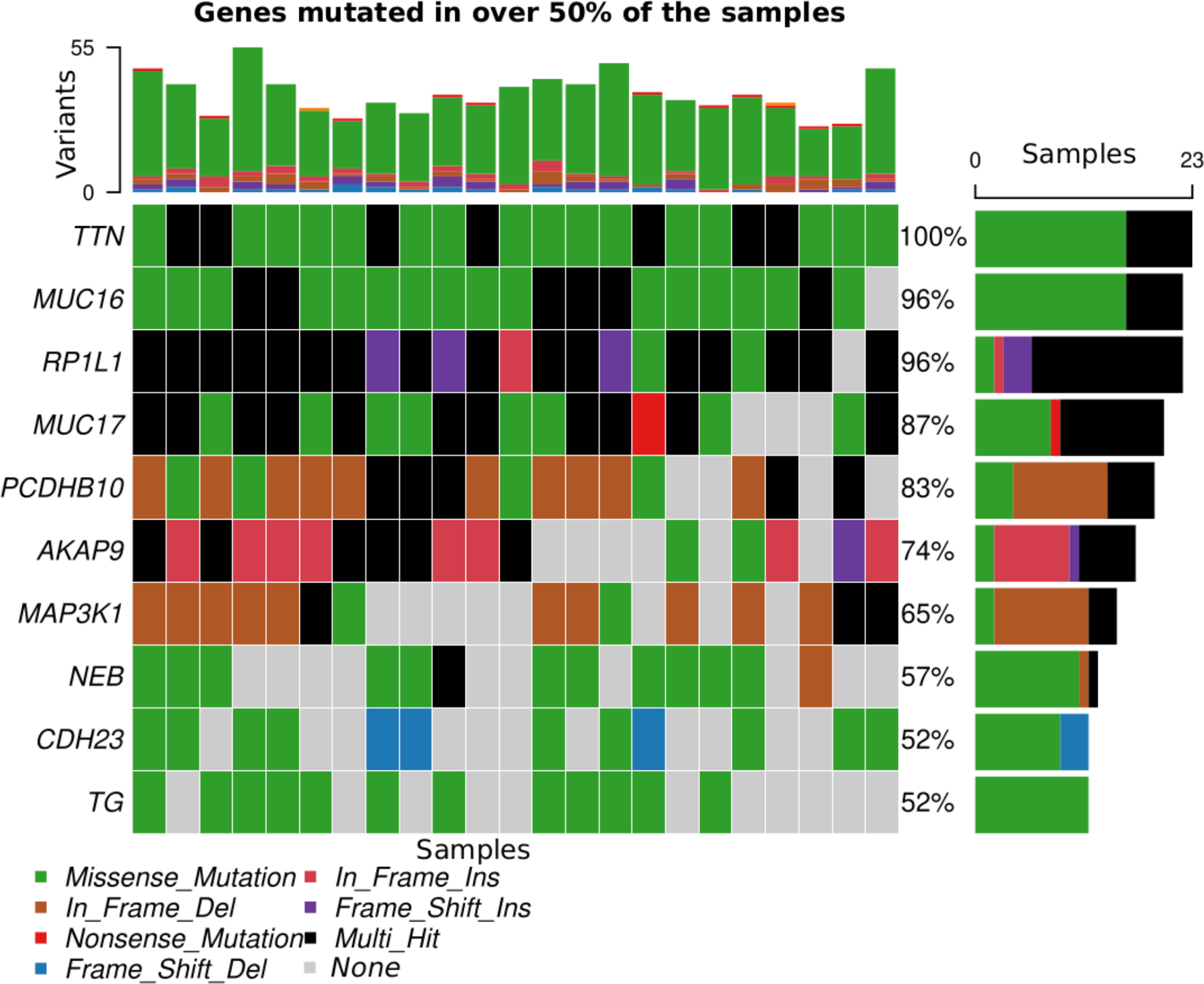
Top 10 genes mutated in >50% of the samples. Each color corresponds to a variant class listed at the bottom of the figure apart from gray, which indicates absence of mutation.

**Figure 4:**
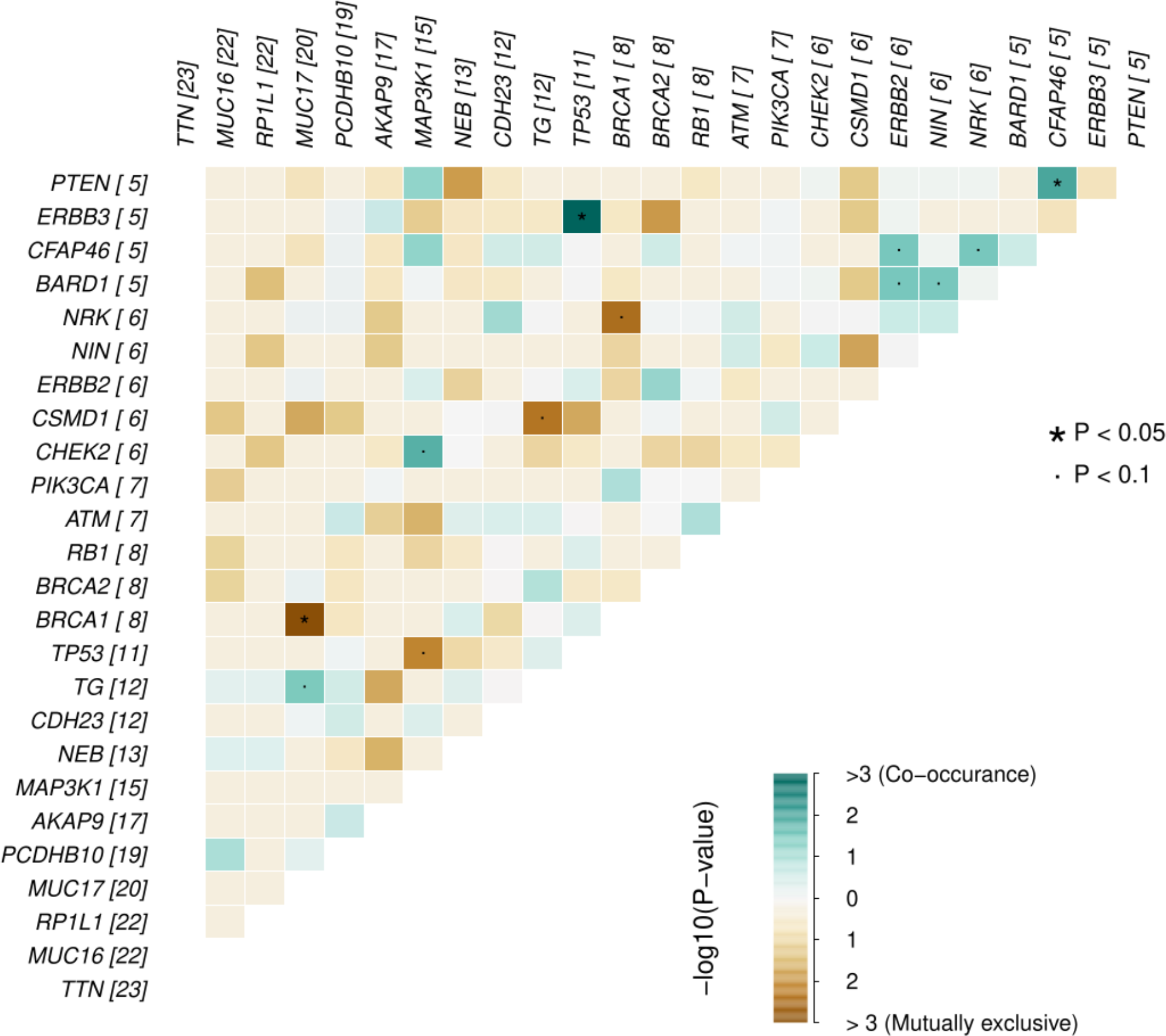
Probability of mutations in any two genes co-occurrence or being mutually exclusive in the breast cancer genes for the 23 Kenyan patients. The numbers in parenthesis alongside each gene represents the number of missense mutations for that gene in the samples.

**Figure 5:**
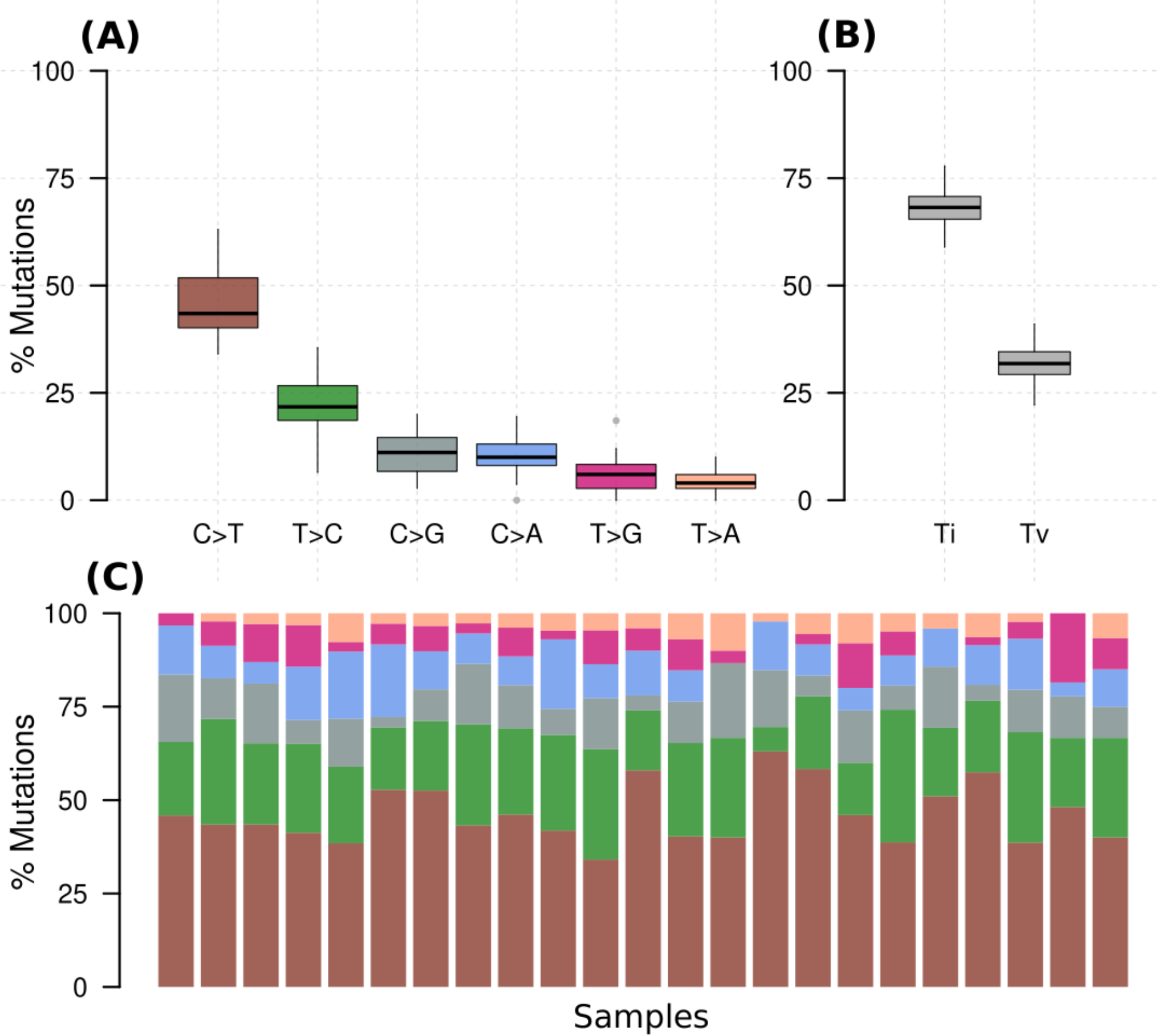
**A**) Percentage of various substitution types in all samples, **B**) percentage of transversions (interchange of purines for pyrimidine) and transition (interchange of either purines or pyrimidines) for all samples, **C**) percentage of the substitutions in each of the samples with colors denoting the various types in A.

### Neoantigen burden

In an analysis that included all the genes (10260), an average of 1465 neoantigens had a ≤500nM median IC50 binding score and >1 TPM expression level in any of the 23 patients and their presence significantly correlated with the somatic mutations (*R*^2^=0.570, *P*=0.001) (Figure 6). Out of the 62 COSMIC genes that were mutated in the tumor tissue, 58 genes produced at least one neoantigen. After filtering for genes that produced at least two neoantigens, 44 genes had a mean of 10.5 neoantigens ranging from 2 to 93. A total of 477 putative neoantigens were identified in these 44 genes across the 23 patients (Figure 7) predominantly derived from missense mutations (88%), indels (6%) and frameshift mutations (6%) (Figure 8). Most of the neoantigens were produced in the TNBC subtype with an average of 25 neoantigens, followed by HR+/HER2-at 20 neoantigens and HER2+ with an average of 19 neoantigens (Figure S1). Notably, 78% of the putative breast cancer neoantigens were patient-specific (Table S2). HLA-C*06:01 allele was associated with majority of neoantigens (194), followed by HLA-A*30:01 (131), HLA-A*02:01 (103), and HLA-B*58:01 (49). Among the genes of interest that produced putative neoantigens include *MUC17*, *TTN*, *MUC16*, *AKAP9*, *NEB*, *RP1L1*, *CDH23*, *PCDHB10*, *BRCA2*, *TP53*, *TG*, *RB1* among others (Figure 7, Table S3).

**Figure 6:**
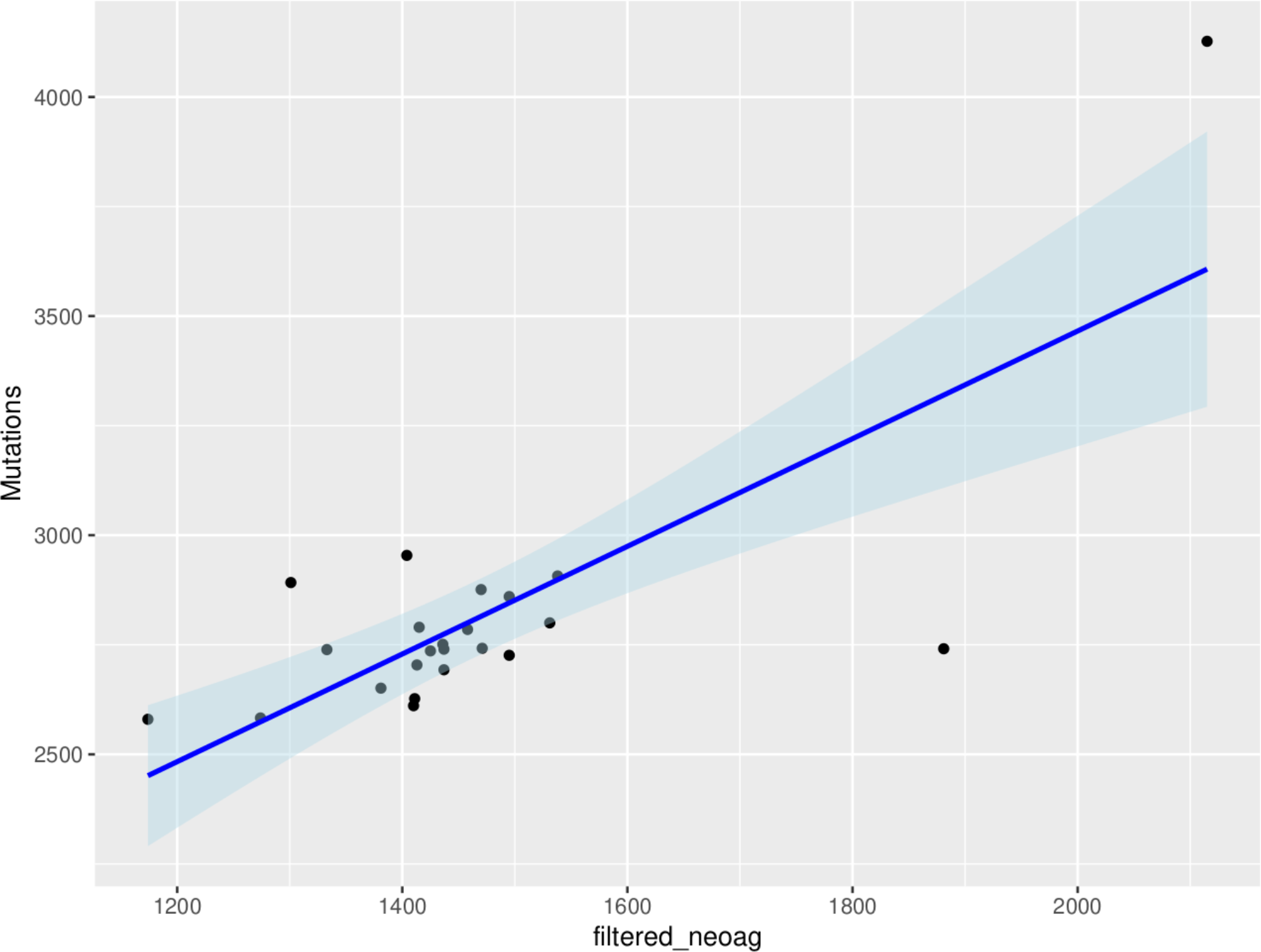
Correlation between tumor mutational burden and neoantigen burden for all the genes in the 23 patients. The neoantigens are filtered for high affinity (IC50 ≤ 500nM) and expression (transcripts per million, TPM>1) in tumor samples.

**Figure 7:**
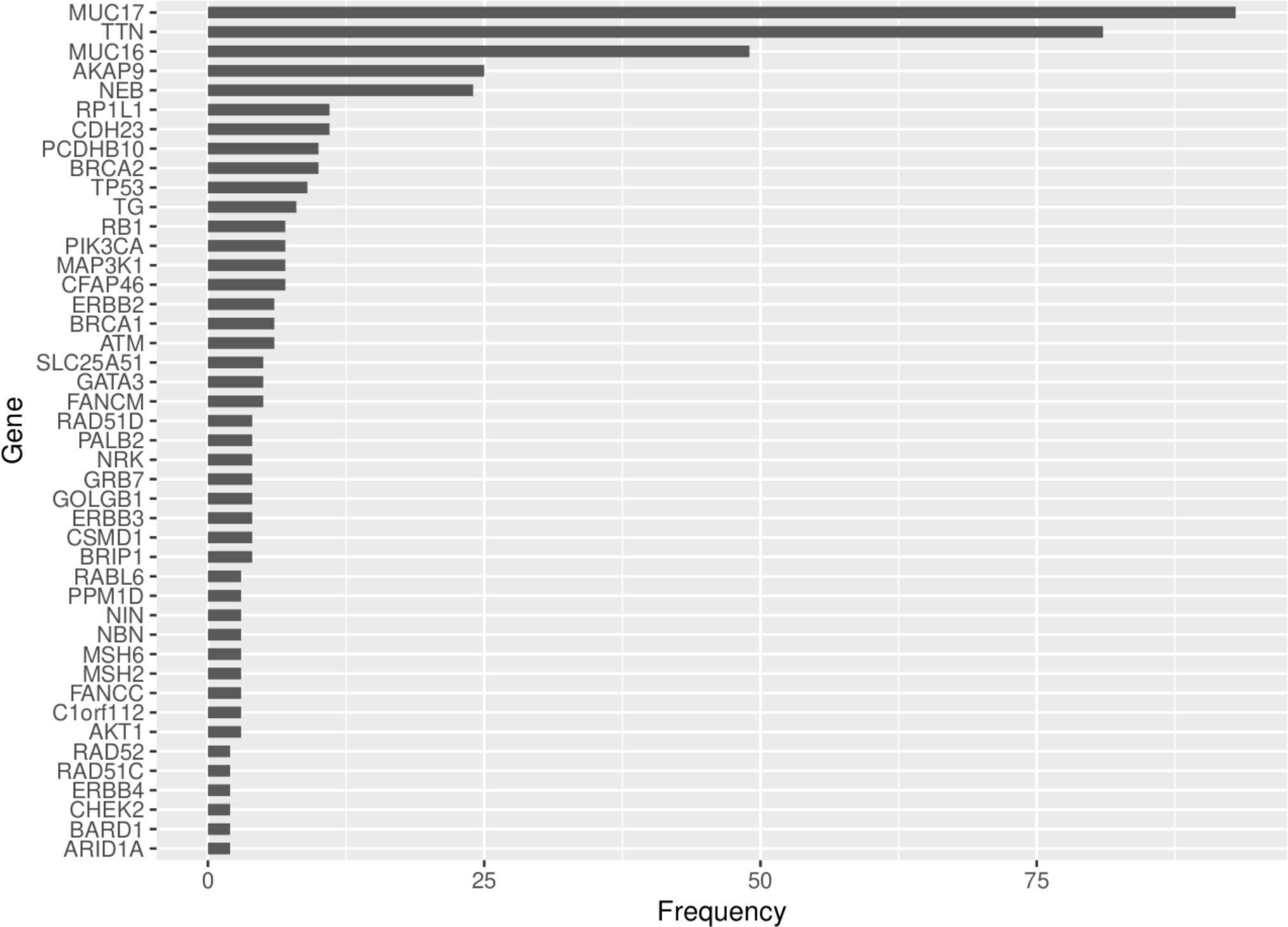
Frequency of neoantigens derived from the COSMIC genes that were mutated in the tumor tissue and produced >1 neoantigens for the 23 patients.

**Figure 8:**
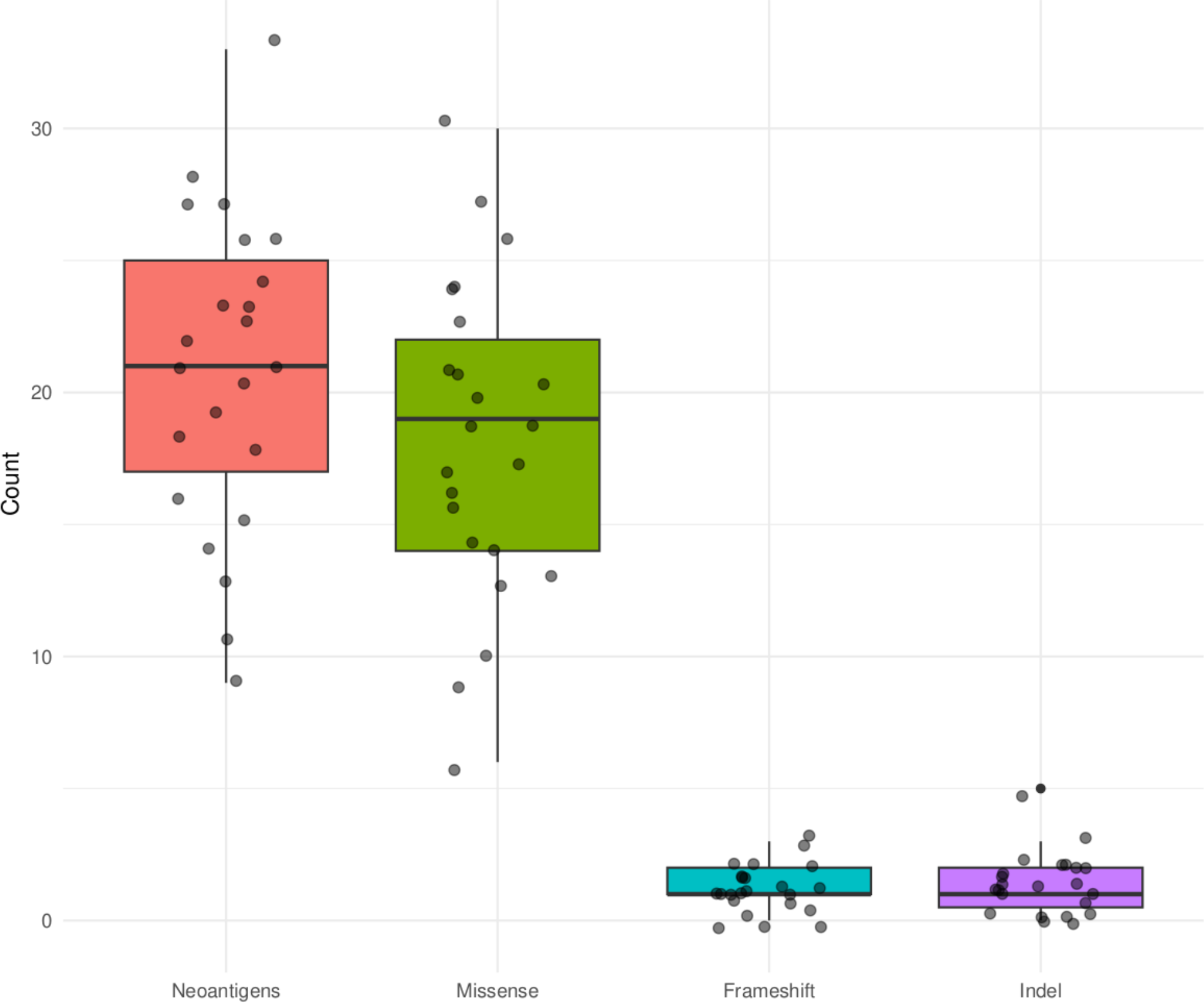
Summary of mutation types that produced putative neoantigens for the COSMIC genes that were mutated in the tumor tissue in the 23 Kenyan patients.

## DISCUSSION

We analyzed the mutational burden and predicted the neoantigen repertoire in 23 Kenyan breast cancer patients using WES and RNA sequencing data. Among the different breast cancer subtypes, we found that the TNBC molecular subtype had the highest mutational and neoantigen burden although there was no significant difference among the subtypes (Figure S1, Table S4). This is consistent with other studies (Narang et a., 2019). TNBC origin is not well understood although it is reported to be heterogeneous in nature relying on different signaling pathways such as JAK/STAT, PI3K/AKT/mTOR or NOTCH, cell cycle regulators (*TP53*) and genome integrity genes (*BRCA1/2*) (Benvenuto et al., 2019). This makes it a disease that is difficult to manage because we do not have a clear understanding of the molecular mechanisms driving it. Yet, the high mutational and neoantigens burden combined with the patient specificity may provide an untapped opportunity to design and optimize personalized immunotherapy for this subtype.

In contrast to most populations where *TP53*, *PIK3CA* and *GATA3* are the most mutated genes (Pan et al., 2020; Pipek et al., 2023; Tang et al., 2023), in our study population, three genes *MUC16*, *MUC17* and *TTN* were highly mutated in over 50% of the samples and produced the highest number of neoantigens. *MUC16* has been reported to take part in breast cancer progression and metastasis when overexpressed due to its influence on cell cycle and survival through the JAK2/STAT3 pathway (Lakshmanan et al., 2012). It has been reported as one of the highly mutated genes in breast cancer (Wang & Guda, 2016). *MUC16* has also been described as a marker for disease progression, recurrence, and chemotherapy response (Felder et al., 2014). A high mutation frequency for *MUC17* and *TTN* have recently been reported as an unexpected finding in a study of early onset breast cancer (EOBC) in Taiwanese women (Midha et al., 2020). *MUC17* may influence chemoresistance and has recently been reported as a driver gene in adult gliomas (Al Amri et al., 2020; Machado & Ferrer, 2023). For *TTN*, Oh et al. (2020) found that mutations in *TTN* correlate with tumor mutational burden and high microsatellite instability, which is associated with poor breast cancer prognosis. Thus, the role of *MUC17* and *TTN* should further be investigated on how mutations in them may relate to early onset of breast cancer in Kenyan patients (Tang et al., 2023).

We found that *TP53* gene mutations significantly co-occurred with *ERBB3* mutations and so did mutations in *PTEN* and *CFAP46*, whereas *BRCA1* and *MUC17* mutations never co-occurred. *TP53* mutations are associated with tumor aggression and are found in about half of HER2-amplified tumors (Marvalim et al., 2023). The *TP53* mutations have been implicated in poor prognosis of HER2+ subtypes compared to other subtypes (Dumay et al., 2013). *PTEN* is a tumor suppressor gene, whose mutation has been associated with initiation, progression, and metastasis of breast cancer (Chen et al., 2022). On the other hand, although *CFAP46* role in breast cancer is not yet clear, gene fusion involving various other genes such as *VTI1A* (reported to cause the initiation of glioma and other cancers) has been reported to play a role in breast cancer (Tsuge et al., 2019).

Breast tumors with either germline or somatic *BRCA1* mutations show no difference in their cancer biology, but inherited mutations in this gene confers a very high lifetime risk of developing breast cancer (Milne & Antoniou, 2011; den Brok et al., 2017; Bodily et al., 2020). This could be the reason such mutations do not necessarily need to co-occur with other gene mutations to initiate or promote breast cancer progression. In our study, *BRCA1* was not among the highly mutated genes considering all mutations but was among the genes with high number of missense mutations (Figure 4). In contrast, *MUC17* mutations were among the most prevalent. Given the role of *MUC17* mutations in chemoresistance and in early onset breast cancer (Al Amri et al., 2020; Machado & Ferrer, 2023), its high prevalence and exclusive occurrence in the Kenyan samples that are prone to early onset of breast cancer should be investigated further.

Similar to most studies on neoantigen prediction in breast cancer, we have found that neoantigens burden is positively correlated with tumor mutational burden and that neoantigens were patient-specific (Narang et al., 2019; Animesh et al., 2022). Although most of the top 10 mutated genes (80%) were also the top 10 in the number of neoantigens generated, genes like *TP53* and *PIK3CA* that are reported to be highly mutated in most patient cohorts were not among the top 10 mutated genes in this study, but generated among the highest number of neoantigens (Figure 6; Figure 7). *ARID1A* gene, which showed unique mutational profile in Kenyan population using exome data compared to African American and Asian population (Tang et al., 2023), was not among the highly mutated, but produced neoantigens. We found that most neoantigens were derived predominantly from missense mutations (88%), compared to indels and frameshift mutations (12%). This is consistent with other studies although the majority do not predict neoantigens from indels and frameshift mutations (Morisaki et al., 2021). Similar to other studies, the TNBC subtype had more neoantigens, compared to HR+/HER2- and HER2+ subtypes (Narang et al., 2019; Morisaki et al., 2021).

In our small sample cohort, we have been able to identify putative neoantigens that show patient-specificity and thus are important in tailored treatment. Interestingly, the mutations and neoantigens in this population are predominantly derived from a unique set of genes (*MUC16*, *MUC17*, *TNT*) compared to other populations, which provide an opportunity for validation in a much larger sample cohort. We predicted neoantigens based on binding affinity to HLA class I only as it is the most important class of antigen binding proteins in cancer immunity. However, HLA class II-based neoantigens may also have a role in tumor immune response (Alspach et al., 2019). Moreover, we did not investigate the expression of the predicted neoantigens on tumor cells alongside the MHC class I molecules and their ability to activate T cells. This being a discovery study, validation of the findings need to be done in a larger cohort while addressing the highlighted limitations of this study.

Taken together, our findings corroborate the neoantigen profile in breast cancer, highlighting the patient specificity in Kenyan population breast cancer mutational and neoantigens signatures. We also describe putative neoantigens that could be used as markers for breast cancer diagnosis, treatment monitoring, and development of novel immunotherapy.

## Acknowledgment

We would like to thank the patients for their consent to provide samples and Aga Khan University Hospital (Nairobi) and AIC Kijabe Hospital (Kijabe) for granting access to patient samples.

## Data accessibility

WES data is accessible at SRA database Accession number: PRJNA913947, while RNA-seq data is accessible at the GEO database under Accession number: GSE225846. All other datasets for this study are included in the article’s Supplementary Material.

## Authors’ contribution

FM conceived the idea and designed the research project, collected the samples and assembled experiment materials. GW, JG, KM and MM performed the data analysis. FM and GW wrote the manuscript. ARH, SS, SA helped with drafting and reviewing the manuscript. All authors contributed to the revision and final editing of the manuscript prior to submission.

## Supplementary Materials

**Figure S1:**
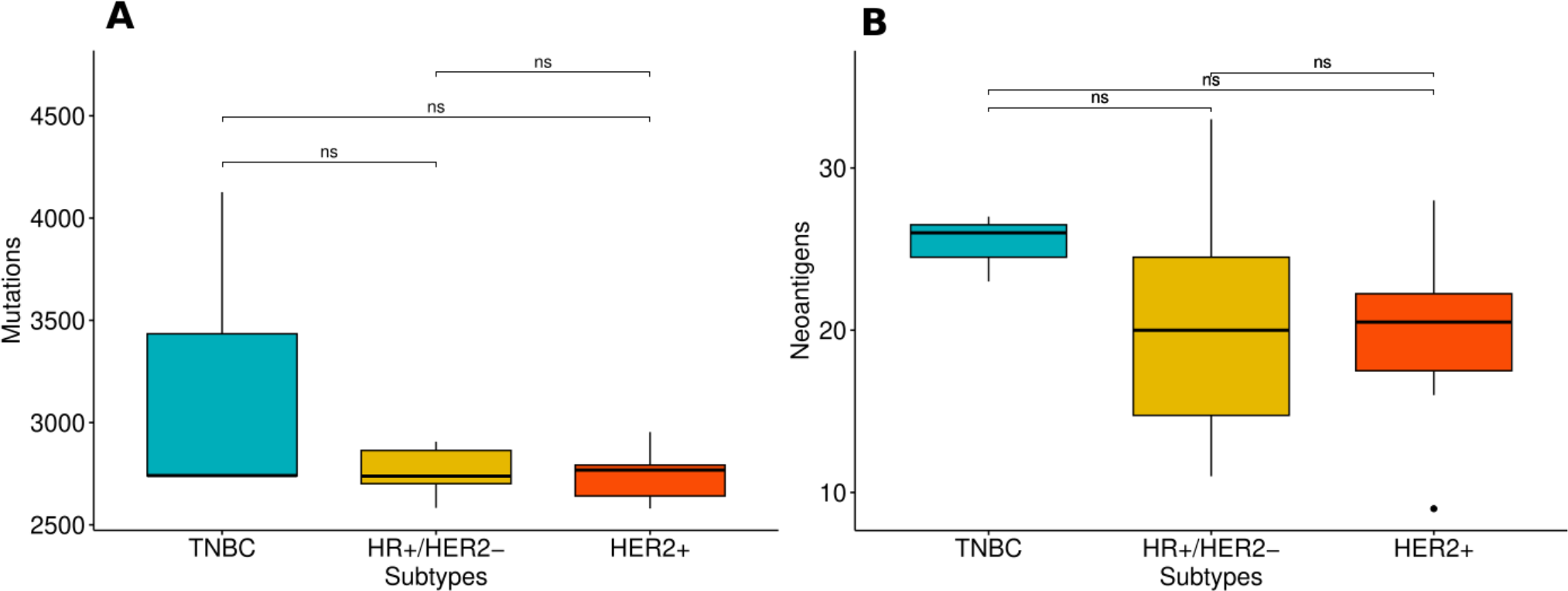
Statistical pairwise test (Wilcoxon’s test) for differences in mutational burden (A) and neoantigens counts (B) for the 23 samples.

**Table S1:** Sample characteristics of the 23 Kenyan patients used in this study.

**Table S2:** Putative neoantigens for each of the 23 Kenyan patients. Cells in red indicate that the neoantigen is shared by at least 2 patients.

**Table S3:** Summary of the roles of the top ten genes that generated a high number of neoantigens.

**Table S4:** Summary of total mutations and proportion of mutation types, total neoantigens, filtered total neoantigens (filtered for high affinity (IC50 ≤ 500nM) and expression [transcripts per million, TPM>1] in tumor samples) and filtered putative neoantigens from COSMIC 44 genes mutated in tumor tissue, and proportion of mutation types that generated them per breast cancer subtype for the 23 samples

